# Association between Non-Dietary Cardiovascular Health and Expenditures Related to Acute Coronary Syndrome in the US between 2008–2018

**DOI:** 10.1101/2023.04.28.23289253

**Authors:** Abraham Enyeji, Noël C. Barengo, Boubakari Ibrahimou, Gilbert Ramirez, Alejandro Arrieta

## Abstract

**Background:** Acute Coronary Syndrome (ACS) causes the most deaths in the United States and accounts for the highest amount of healthcare spending. Cardiovascular Health (CVH) metrics have been widely used in primary prevention, but their benefits in secondary prevention on total healthcare expenditures related to ACS are largely unknown. This study aims to quantify the potential significance of ideal CVH scores as a tool in secondary cardiovascular disease prevention.

**Methods:** In a cross-sectional analytical study, ten years of Medical Expenditure Panel Survey (MEPS) data from 2008 to 2018 were pooled, comparing ACS to non-ACS subgroups, utilizing a Two-part model with log link and gamma distribution, since our sample had both positive and zero costs. Conditional on positive expenditure, healthcare expenditure amounts were measured as a function of ACS status, socio-demographics, and CVH while controlling for relevant covariates. Finally, interactions of ACS with CVH metrics and other key variables were included to allow for variations in the effect of these variables on the two subgroups.

**Results:** Improvements in CVH scores tended to reduce annual expenditures to a greater degree percentage-wise among ACS subjects compared to non-ACS groups, even though subjects with an ACS diagnosis tended to have approximately twice as big expenditures as similar subjects without an ACS diagnosis. Meanwhile, the financial impact of an ACS event on total expenditure would be approximately $88,500 ([95% CI, $70,200–106,900; *p* < 0.001]), and a unit improvement in CVH management score would generate savings of approximately $4160 ([95% CI, $5390–2950; *p* < 0.001]) in total health expenditures.

**Conclusion:** Effective secondary preventive measures through targeted behavioral endeavors and improved health factors, especially the normalization of hypertension, diabetes mellitus, body mass index, and smoking cessation, have the potential to reduce medical spending for ACS subgroups.

## 1. Introduction

Every year, approximately 635,000 Americans have a new episode of Acute Coronary Syndrome (ACS), and approximately 280,000 have a recurrent event [1,2]. In more recent years, the cost of index hospitalization due to ACS averaged $18,931 and the annual 30-day hospital cost of invasive procedures related to ACS is estimated at $10.8 billion [3]. In terms of direct medical expenditures, ACS costs Americans more than US $150 billion annually, with approximately 60–75% of these costs related to primary inpatient care and recurrent ACS events or readmissions [4].

The term ACS refers to a spectrum of conditions in which myocardial ischemia or infarction develops due to the acute occlusion of coronary blood flow to any part of the heart. The symptoms of ACS can include chest pain or discomfort, shortness of breath, nausea, sweating, and lightheadedness. There are three main types of ACS (see Table A1 in Appendix A): Unstable angina, non-ST segment elevation myocardial infarction (NSTEMI), and ST-segment elevation myocardial infarction (STEMI) [5]. The most common cause of ACS is atherosclerosis, a condition in which plaque builds up in the arteries that supply the heart with blood. The plaque build-up can cause a partial or complete blockage of the involved artery, which results in reduced blood flow to the heart.

For reasons that are not fully understood, pro-thrombotic activation prevails over the healing response in some patients, causing the equilibrium to shift towards a flow-limiting thrombus and clinical manifestation of ACS. This pathophysiological evidence suggests reasons why patients with a history of ACS are more susceptible to a recurrent ACS event, because of their biological predisposition to develop atherothrombotic plaques. It, therefore, seems as though the likelihood of having a recurrent ACS event rises with every ACS event acquired due to biological predisposition in the tissue of ACS patients [5], hence the exponential rise in costs for ACS treatment.

While not directly comparable due to differences in methodology, the annual cost of ACS in 2016 was $150 billion [6] lower than other chronic diseases such as obesity ($260 billion) [7] and cancer ($157 billion) [8]. While calculations of the costs of obesity often involve the costs associated with the treatment of obesity-related conditions such as type 2 diabetes, heart disease, and stroke, there is generally no direct relationship between the expenditures of ACS and cancer, as these are two different medical conditions that require different approaches to diagnosis, treatment, and prevention.

Previous researchers have shown that routine assessment of Cardiovascular Health (CVH) scores as a tool for primary prevention would ease the identification of individuals at risk of Cardiovascular Disease (CVD) [9]. Achieving an ideal CVH status would also significantly lower total medical expenditure [10]. The American Heart Association’s (AHA) Life’s Simple 7 (LS7) is an accurate determinant for CVH [11] because achieving ideal CVH scores in middle age is linked to improved outcome prognosis after a Myocardial infarction event in later life. Moreover, secondary preventative measures that utilize CVH scores have a significant impact on reducing the incidence of heart failure [12]. These findings, therefore, suggest a benefit in secondary prevention for having improved CVH status in midlife [10]. However, it is unknown whether achieving ideal levels of CVH management scores as a tool for secondary prevention is associated with increased total healthcare savings for individuals with ACS.

A few studies have successfully demonstrated the benefit of healthcare savings with improved CVH scores as a measure of primary prevention [13,14]. Therefore, to quantify the potential significance of ideal CVH scores in secondary prevention, we examined data from the Medical Expenditure Panel Survey (MEPS) database to explore the impact of improved CVH scores on total healthcare savings among individuals with ACS. This study aims to analyze the association between CVH and the economic burden and enhancement of secondary prevention in individuals aged nineteen and above who have demonstrated ACS. Considering the substantial economic effects of ACS, this cross-sectional study will analyze the effects of adherence to the AHA’s construct on healthcare expenditures in a socioeconomically diverse cohort of patients with and without ACS.

CVH assessment is achieved by a set of criteria consisting of the cumulative effect of modifiable risk factors. These include a measurable set of seven modifiable risk factors or behaviors that prevent the incidence of ACS and are referred to by the AHA as the CVH metrics [15,16]. These metrics are used by the AHA as a standard for the assessment of CVD risk. The elements of CVH are often used to identify and quantify known risk factors for CVD, such as smoking, physical inactivity, body mass index (BMI), diet, blood pressure, cholesterol, and glucose levels. These CVH components are the factors research has shown can be modified to reduce the risk of ACS through lifestyle or behavioral changes, especially when one is younger [17]. These CVH components make up the building blocks for the CVH metric assessment and consist of four health behaviors: (1) Dieting, (2) physical activity, (3) smoking, and (4) maintaining the right BMI; and the three health factors are (1) normalizing blood cholesterol, (2) blood pressure, and (3) blood glucose levels. For this research, the term Non-dietary CVH is used instead of CVH because the self-administered questionnaires do not address dieting.

## 2. Materials and Methods

### 2.1. Study Design

This study used pooled cross-sectional, secondary data from the Medical Expenditure Panel Survey (MEPS) database from 2008 to 2018. The MEPS is a set of large-scale national surveys of households, individuals, insurance companies, health providers (doctors, pharmacies, and hospitals), and employers across the U.S., sponsored by the Agency for Healthcare Research and Quality (AHRQ). The panel consists of randomly sampled noninstitutionalized U.S. civilians providing nationally representative estimates of sociodemographic characteristics, medical conditions, and utilization and costs [18].

Numerous studies have proven the validity of the Medical Expenditure Panel Survey (MEPS) in terms of healthcare utilization and expenditure. In some validation studies of household reports of healthcare utilization in the MEPS, researchers found that respondents in the validation sample accurately report inpatient hospitalizations. Even though emergency departments and office visits were underreported, behavioral analyses remain largely unaffected because underreporting cuts across all sociodemographic groups, and all groups of Medicare beneficiaries in the matched sample were uniformly affected [19,20].

### 2.2. Study Population

Adults aged 19 and above, with and without ACS, who participated in the Medical Expenditure Panel Survey (MEPS) between 2008 and 2018 were included. To obtain the final sample, three MEPS components emerged: (1) The household component, which includes data on demographic characteristics, charges, and payments; (2) the events component, which includes visits related to any specific health condition; and (3) the medical conditions component, which includes a “current” condition per person at any time during the data year. The data files were merged using the conditions event link file for each year. Pooling the merged data for 10 years from 2008 to 2018 allowed for a larger analyzable population of adults with and without ACS. The year 2017 was excluded because the variable for body-mass index (BMI), an essential component of the CVH metrics, could not be created for that year. A looping algorithm was used to rename all cost categories so that all different types of medical costs could be pooled to generate a total expenditure variable for an individual for a specified year of MEPS data. The cost types analyzed in MEPS include inpatient, outpatient, office-based, drug-prescription, home health, and emergency room event expenses.

Individuals with ACS were identified for the years 2008–2015 using the International Statistical Classification of Diseases, Clinical Modification ICD-9 CM diagnosis of the condition (410, 411, 412, 413, 414), and ICD-10 CM (I20, I21, I22, I23, I24) for the years 2016 and 2018. Charlson’s Comorbidity Index was generated using the same ICD-9 and ICD-10 CM codes used to specify medical conditions. ACS refers to the self-reported history of diagnosis of unstable angina and acute myocardial infarction (AMI), ST-elevation myocardial infarction (STEMI), or Non-ST elevation myocardial infarction (NSTEMI), acute and subacute forms of ischemic heart disease. The study population was limited to noninstitutionalized US adults with or without established ACS who were ≥19 years.

### 2.3. Primary Outcome

The primary dependent variable was the total healthcare expenditure. This variable was generated from the sum of service categories that included inpatient, outpatient, home health, office-based, drug prescription, and emergency room visits. MEPS collects this information from medical providers, insurance companies, employers, and hospitals.

### 2.4. Measuring Non-Dietary Cardiovascular Health

For this study, the primary independent variable of interest was the number of achieved CVH scores for each participant, represented by the CVH composite score, with a range of 0 to 6. The non-dietary LS7 components examined in this study included inadequate physical activity, obesity, smoking, hypercholesterolemia, hypertension, and diabetes mellitus. Diet was not assessed in MEPS, and therefore, the maximal score of 6 was achieved as a composite score, as occurred in other research [21]. Participant responses from the self-administered questionnaire were utilized to determine the CVH status of participants, classifying everyone with a binary variable (favorable [1] versus unfavorable [0]). Those who reported a diagnosis of a cholesterol disorder, hypertension, or diabetes mellitus, smoked within less than one year of the time of the interview, did not engage in moderate vigorous physical activity 5 times a week, or had a BMI >25 or <18 kg/m^2^ were classified as having an unfavorable risk factor. They were then assigned a 0, while those who did not exhibit any of these adverse pre-conditions were assigned a value of 1. The sum of values assigned to determine a participant’s CVH score ran from 0 through 6.

Individual comorbidity levels were assessed using the Grouped Charlson Comorbidity Index (GCCI), which is a method of categorizing comorbidities of patients based on the International Classification of Diseases (ICD) diagnosis codes found in administrative data, such as hospital abstract data [22]. It stratifies comorbid conditions by disease severity using associated weights (from 0 to 6) based on the adjusted risk of mortality or resource use [23].

### 2.5. Covariates

Education, age, insurance, sex, race/ethnicity, and family income were examined as factors that tend to be associated with healthcare expenditures and therefore were examined as covariates in the determination of the association between CVH and healthcare expenditures overall and in ACS cohorts. Participants’ sex was classified as self-reported male and self-reported female. Self-reported races or ethnicities were defined as White (non-Hispanic), Black (non-Hispanic), Hispanic, and Asian (non-Hispanic). Furthermore, self-reported family income level (poor or very low income: <125% of the federal poverty level; low income: 125% to <200% of the federal poverty level; middle income: 200% to <400% of the federal poverty level; or high income: ≥400% of the federal poverty level) and educational level (having less than a bachelor’s degree, having achieved an equivalent of a bachelor’s or master’s degree, and then a Doctoral or professional degree) were defined. Health insurance status was divided into private, public (Medicare and Medicaid), and uninsured.

### 2.6. Statistical Analysis

Descriptive statistics to summarize frequency distributions were used with corresponding weighted proportions by ACS versus non-ACS status. Since the sample had both positive and a few negative or zero costs (approximately 4% of our sample) for some participants, a two-part econometric model was used with log link and gamma distribution. This essentially determined the likelihood of having a positive expenditure, addressing a potential selection effect bias. The same specifications were used for both the Probit and Generalized linear Model (GLM) stages, prioritizing significance in the second stage and controlling for sex, race/ethnicity, age, level of income, education levels, health insurance, and the Charlson Comorbidity Index, used to explore the associations of CVH status with total healthcare expenditure.

ACS was interacted with CVH metrics, age, and highest educational levels attained to be flexible about the impact of ACS on expenditures, allowing variations in the effects of these variables to reflected across the ACS subgroups. In the final step, the CVH metrics were replaced with all CVH components, and the two-part model regression was repeated in order to capture the association between LS7 components with total costs after controls. Cells contained coefficients and two-sided p values and *s for whether a coefficient was statistically significant with p less than 0.05. In the two-part model, a Probit model was used in the first stage to assess the likelihood of positive expenditure on healthcare and a generalized linear model (GLM) with log link and Gamma distributions in the second to determine the expected total healthcare expenditure. The two-part model used is explained below:

To estimate the total expenditure for patients with and without ACS, the following regression model was:

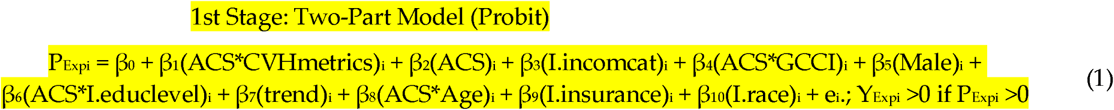

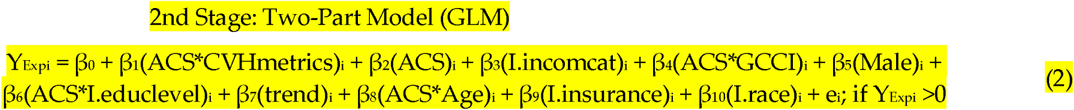

where,

- P_Expi_: represents the likelihood of healthcare expenditure.
- CVHmetrics_i_: represents CVH metrics.
- Age_i_ represents the patient’s age.
- I.incomcat_i_ represents individuals’ income category based on the Federal poverty level.
- GCCI_i_: Charlson’s Comorbidity Index.
- Male_i_: 1 if the subject self-identifies as male, 0 otherwise.
- I.educlevel_i_: indicates the level of highest education attained.
- ACSi: ACS indicator.
- trend_i_: represents the effect of trend.
- I.insurance_i_: represents categorical variable for insurance with three categories.
- I.race_i_: represents the impact of race utilized as a categorical variable.
- Y_Expi_ represents the total annual healthcare expenditure.
- e_i_: Error term with normal distribution (0 = mean, 1 = SD)

All analyses were conducted using Stata version 17 (StataCorp LLC, College Station, TX, USA), adjusting for MEPS complex survey design. The study received an exemption from the Institutional Review Board (IRB Protocol Exemption Number: IRB-22-0170).

## 3. Results

### 3.1. Characteristics of the Sample

The study population consisted of 90,732 individuals from 2008–2018. They were individuals 19 years old or older with a BMI ranging between 18 and 25 kg/m^2^, for normal BMI, and who either had or did not have ACS. The mean age was 67.2 years (CI: 66.82–67.5) for the ACS subgroup and 48.9 years (CI: 48.7–49.0) for the non-ACS sub-groups, and 56.4% were female. The distribution of demographic characteristics for the study population is shown in Table 1 below.

**Table 1.** Summary statistics.

| Prevalence, % (95% CI) | ACS | Non-ACS | Diff (*p*-Values) | Combined Sample |
| --- | --- | --- | --- | --- |
| **Outcomes** |  |  |  |  |
| Health Expenditures = 0 | 1.7 (1.6–2.3) | 3.8 (3.6–3.9) | *p* < 0.001 | 3.7 (3.5–3.8) |
| Average Healthcare Expenditures if >0 (\$) | 177,657 (168,339–186,974) | 46,981 (45,962–47,999) | *p* < 0.001 | 54,498 (53,382–55,615) |
| **CVH Components** |  |  |  |  |
| Inadeq. Phy. Activity | 47.7 (46.4–49.0) | 56.1 (55.8–56.5) | *p* < 0.001 | 55.7 (55.3–56.0) |
| High Cholesterol | 80.2 (79.1–81.3) | 37.7 (37.3–38.0) | *p* < 0.001 | 40.1 (39.8–40.5) |
| Hypertension | 85.3 (84.4–86.3) | 37.7 (41.7–42.4) | *p* < 0.001 | 44.5 (44.2–44.9) |
| Diabetes Mellitus | 39.9 (38.5–41.2) | 14.7 (14.5–14.9) | *p* < 0.001 | 16.1 (15.9–16.3) |
| Normal BMI (18–25) | 30.3 (29.1–31.6) | 33.3 (33.0–33.6) | *p* < 0.001 | 33.1 (32.8–33.5) |
| Smokers | 19.8 (8.7–20.8) | 17.4 (17.1–17.6) | *p* < 0.001 | 17.5 (17.3–17.8) |
| **Self-identified gender** |  |  |  |  |
| Male | 59.7 (58.4–61.0) | 42.9 (42.6–43.2) | *p* < 0.001 | 43.9 (43.6–44.2) |
| **Mean Age** |  |  |  |  |
| Age | 67.1 (66.8–67.5) | 48.8 (48.6–49.0) | *p* < 0.001 | 49.8 (49.7–50.0) |
| **Race/Ethnicity** |  |  |  |  |
| White | 60.4 (59.1–61.7) | 51.1 (50.8–51.5) | *p* < 0.001 | 51.7 (51.3–52.0) |
| Black | 17.7 (16.7–18.8) | 17.9 (17.6–18.2) | 0.7812 | 17.9 (17.6–18.2) |
| Hispanic | 15.2 (14.3–16.7) | 22.5 (22.2–22.8) | *p* < 0.001 | 22.1 (21.8–22.3) |
| Asian | 4.1 (3.6–4.7) | 6.1 (6.0–6.3) | *p* < 0.001 | 6.0 (5.9–6.2) |
| Multi Racial | 1.8 (1.4–2.2) | 1.9 (1.8–2.0) | 0.6721 | 1.9 (1.8–2.0) |
| Prevalence, % (95% CI) | ACS | non-ACS | Diff (*p*-values) | Combined Sample |
| **Household Income (Federal poverty level [FPL])** |  |  |  |  |
| <125% | 18.3 (17.3–19.4) | 18.3 (17.9–18.5) | 0.8017 | 18.2 (17.3–19.4) |
| 125–200% | 15.9 (14.9–16.9) | 14.1 (13.9–14.3) | *p* < 0.001 | 14.2 (14.0–14.5) |
| 200–400% | 30.5 (29.4–31.9) | 29.4 (29.1–29) | 0.0609 | 29.5 (29.2–29.9) |
| >400% | 35.0 (33.7–36.2) | 38.1 (37.8–38.4) | *p* < 0.001 | 37.9 (37.6–38.3) |
| **GCCI (Grouped Charlson's Comorbidity Index)** |  |  |  |  |
| Average GCCI: 0–6 | 3.8 (3.6–3.9) | 1.4 (1.3–1.5) | *p* < 0.001 | 1.5 (1.4–1.6) |
| GCCI = 0 | 67.5 (66.1–68.7) | 88.5 (88.0–89.0) | *p* < 0.001 | 87.3 (87.3–87.8) |
| GCCI = 1 | 28.2 (27.0–29.1) | 9.4 (9.2–9.7) | *p* < 0.001 | 10.8 (10.4–10.7) |
| GCCI = 2 | 3.2 (2.8–3.7) | 1.0 (0.9–1.1) | *p* < 0.001 | 1.2 (1.0–1.22) |
| GCCI = 3 | 0.9 (0.7–1.2) | 0.6 (0.5–0.67) | *p* < 0.001 | 0.6 (0.58–0.59) |
| GCCI = 6 | 0.1 (0.02–0.20) | 0.1 (0.08–0.12) | 0.8751 | 0.1 (0.08–0.12) |
| **Highest attained educational level** |  |  |  |  |
| High school diploma | 72.2 (71.0–73.4) | 58.7 (58.3–59.0) | *p* < 0.001 | 59.5 (59.1–59.8) |
| Bach/Master's degree | 25.8 (24.6–26.9) | 30.1 (29.8–30.5) | *p* < 0.001 | 29.9 (29.6–30.3) |
| PhD/Prof degree | 3.8 (3.3–4.4) | 3.9 (3.7–4.0) | 0.8680 | 3.8 (3.8–4.1) |
| Sample size | 5391 | 88,319 | - | 93,710 |

Means and proportions were compared across all groups. **Abbreviations:** CVH: Cardiovascular Health, BMI: Body Mass Index; ACS: Acute coronary Syndrome; FPL: Federal poverty level; GCCI: Grouped Charlson Comorbidity Index, ranges from 0 to 6 (6: highest comorbidity level; 0: lowest comorbidity level). Confidence Intervals in parenthesis. Alternating

Of the 90,732 adults in 2008–2018, 95.82% did not have an ACS event; 4.1% reported at least one event of ACS. Meanwhile, almost 3.8% of individuals without a history of ACS reported zero expenditure and approximately 1.7% of the ACS population were found to have no expenditure. The average yearly total expenditure for our combined sample was approximately $54,000, compared with approximately $177,000 for the average annual total medical costs of ACS with $47,000 for non-ACS cohorts. Meanwhile, hypertension and high cholesterol prevailed the most amongst ACS groups with 84.9% and 81.2%, respectively, and physical inactivity and hypertension were found to be the most prevalent in the non-ACS groups with 44.9.0% and 35.8%, respectively. These results match the findings of previous researchers [24] (p. 7).

### 3.2. Differences between ACS and Non-ACS Populations

In our analysis, ACS almost perfectly predicts having a positive healthcare expenditure, which is why the first step of predicting whether someone has a healthcare expenditure of zero was only 1.7% based on ACS status (see Table 1 above).

Table 2 sets out the results for the two-stage model that first predicts the probability of positive expenditures and, second, the predicted expenditures on healthcare, with the final column including the marginal impact of a one-unit change in the control variable on the expected expenditures. Having had an ACS event was positively significant for both stages of the model, making it more likely that someone has healthcare expenditure and pays more when they do obtain healthcare. The marginal increase in annual healthcare expenditure amount after an ACS event was $88,560 ([95% CI, $70,200–106,900; p < 0.001]), which was greater in magnitude than most of the other controls included in the two-stage model.

**Table 2.**
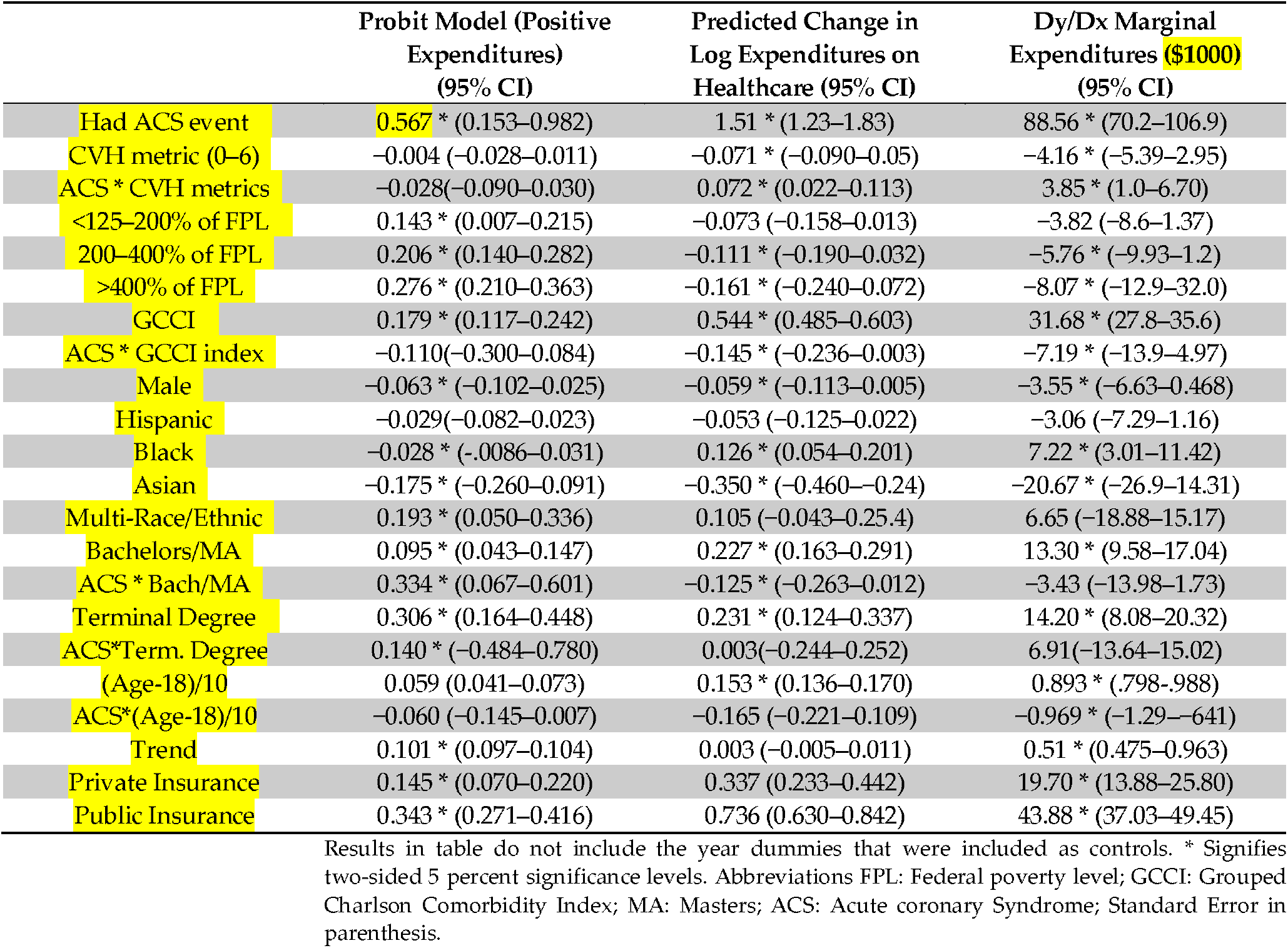
Two-stage model results for the prediction of expenditure based on CVH and ACS status for the years 2008–2018 using the MEPS dataset, showing interaction effects (results in table do not include the year dummies that were included as controls. * Signifies two-sided 5 percent significance levels).

The impact of the CVH metric ranging from 0 to 6, which was included as a linear term, was negative in both stages of the model, suggesting that the impact of rising unit changes in CVH scores would imply reductions in healthcare expenditures. The marginal effect of changing the CVH metric by one unit is a reduction in expected healthcare expenditure by $4160 ([95% CI, $5390–2950; p < 0.001]) per year. If extrapolated, it would imply a reduction in expenditure of $24,960 (p-value < 0.05) if CVH shifted from 0 to 6, slightly more than one-quarter of the magnitude of the impact of having an ACS event. The interaction between having had an ACS event and the CVH metric is more complicated because the interaction effect reinforces the negative effects of a higher CVH metric on the probability of having a positive healthcare expenditure while the positive coefficient for the second stage neutralizes the impact of better cardio-vascular health on the predicted expenditure on healthcare. Which effect proves to be more important? The marginal expenditure in the fourth column of Table 2 suggests that having had an ACS event cancels out the impact of better CVH on reducing expected healthcare expenditure. This suggests that when contrasting the spending differences between persons of different age groups, an increase in age is associated with higher expenditures. However, the improved benefits of being younger leveled out after the onset of ACS. CVH matters before the onset of an ACS event but not afterward.

Some of the other controls have some interesting findings as well. There were three relative income controls based on reported income divided by the federal poverty level. Those with more income were consistently more likely to have positive healthcare expenditure but their expenditure on healthcare tended to be less monetary. The marginal impact is statistically significant and negative for those with more than 2.5 times the federal poverty level, relative to those with less than 1.25 times the federal poverty level in income, while having an income level well above poverty tended to reduce expenditure by almost $8500 per year. The GCCI controlled for the impact of having serious illnesses (or the co-morbidity burden) and tended to raise both the probability of having a positive expenditure and the level of the expenditure. The marginal effect was shy of half the impact of having had an ACS event, or $31,680 ([95% CI, $27,800–35,600; p < 0.001]). The interaction effect between ACS status and GCCI tended to offset some of the impacts of the GCCI but needs to be viewed in conjunction with the predicted impact of an ACS event as shown in the second row of Table 2. The marginal expenditure for the interaction term suggests that the impact of a one-unit-higher GCCI drops from $31,680 to approximately $23,010 if there has been an ACS event.

Participants who self-identified as male relative to all others were both less likely to have healthcare expenditures and tended to spend less on healthcare events compared with those who did not. The marginal effect is a modest $3550 less in healthcare expenditure. This is similar to other race- or ethnicity-related controls. Hispanic participants were less likely to have positive expenditure and to spend less on average with a predicted reduced expenditure of $5300 relative to whites. Blacks had a significantly lower chance of having health expenditure but tended to have higher expenditure on average than whites, with a net increase of $7220 ([95% CI, $3010–11,420; p < 0.05]) per year.

A college education, or greater, predictably statistically significantly increased the predicted level of total healthcare expenditure and tended to increase the probability of having a positive healthcare expenditure. When one goes from obtaining a bachelor’s or a master’s degree, the marginal effect on predicted annual total expenditures is an increase of $13,000 ([95% CI, $9580–17,040; p < 0.001]). When one obtains a terminal degree or Ph.D., the predicted expenditure rises somewhat to be $14,000 ([95% CI, $8080–20,320; p < 0.001]) relative to someone without a bachelor’s degree. The impact of a bachelor’s degree drops less than half if someone has had an ACS event. Thus, prior to an ACS event, one can expect someone to have annual healthcare events and to spend more on healthcare, but after an ACS event, the expected healthcare costs are more comparable between people with or without a bachelor’s degree. The interaction effect of an ACS event with participants who had a final educational degree, such as a PhD, was not statistically significant, which suggests they may still be paying approximately $2500 less than someone with less education than them but are similar otherwise to one who has also had an ACS event.

The results in Tables 2 and A2 (see Appendix A) for health insurance indicate that those with private insurance spent approximately $20,000 ([95% CI, $13,880–25,800; p < 0.001]) more in total annual expenditures compared with the uninsured (Reference), while Medicare/Medicaid-insured individuals spent twice that amount more than the uninsured in annual healthcare.

The age identification variables and their interactions with ACS status consistently show that healthcare expenditures tend to rise in probability and level with age but that the impact of an ACS event is to remove the relative importance of age differences with respect to the probability and level of healthcare expenditures.

### 3.3. Association of Cardiovascular Health with Expenditure

Figure 1 shows that the impact of raising the CVH score on savings is percentage-wise higher for those with ACS compared to non-ACS.

**Figure 1.**
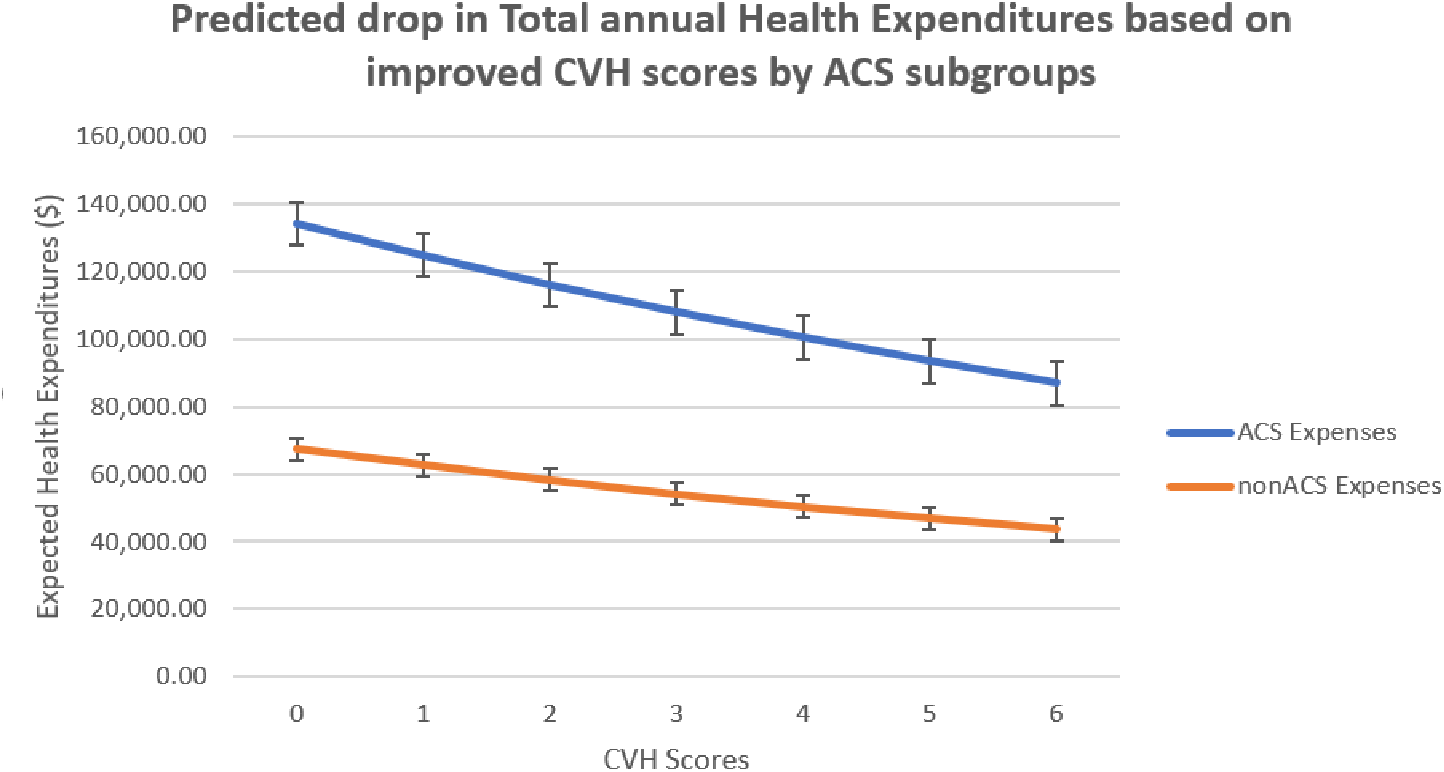
Showing predicted drop in expenditures based on CVH scores by ACS.

Figure 1 shows that improvements in CVH scores predict lower annual healthcare expenditures for both ACS and non-ACS cohorts; however, this is to a greater degree percentage-wise for ACS. These findings are consistent with other researchers who have demonstrated that an improved CVH profile or LS7 score is associated with significantly lower total medical expenditure and healthcare utilization in a large, young, ethnically diverse population [14].

Figure 2 shows the marginal effects, which reflect the negative association between ACS and CVH scores across the board.

**Figure 2.**
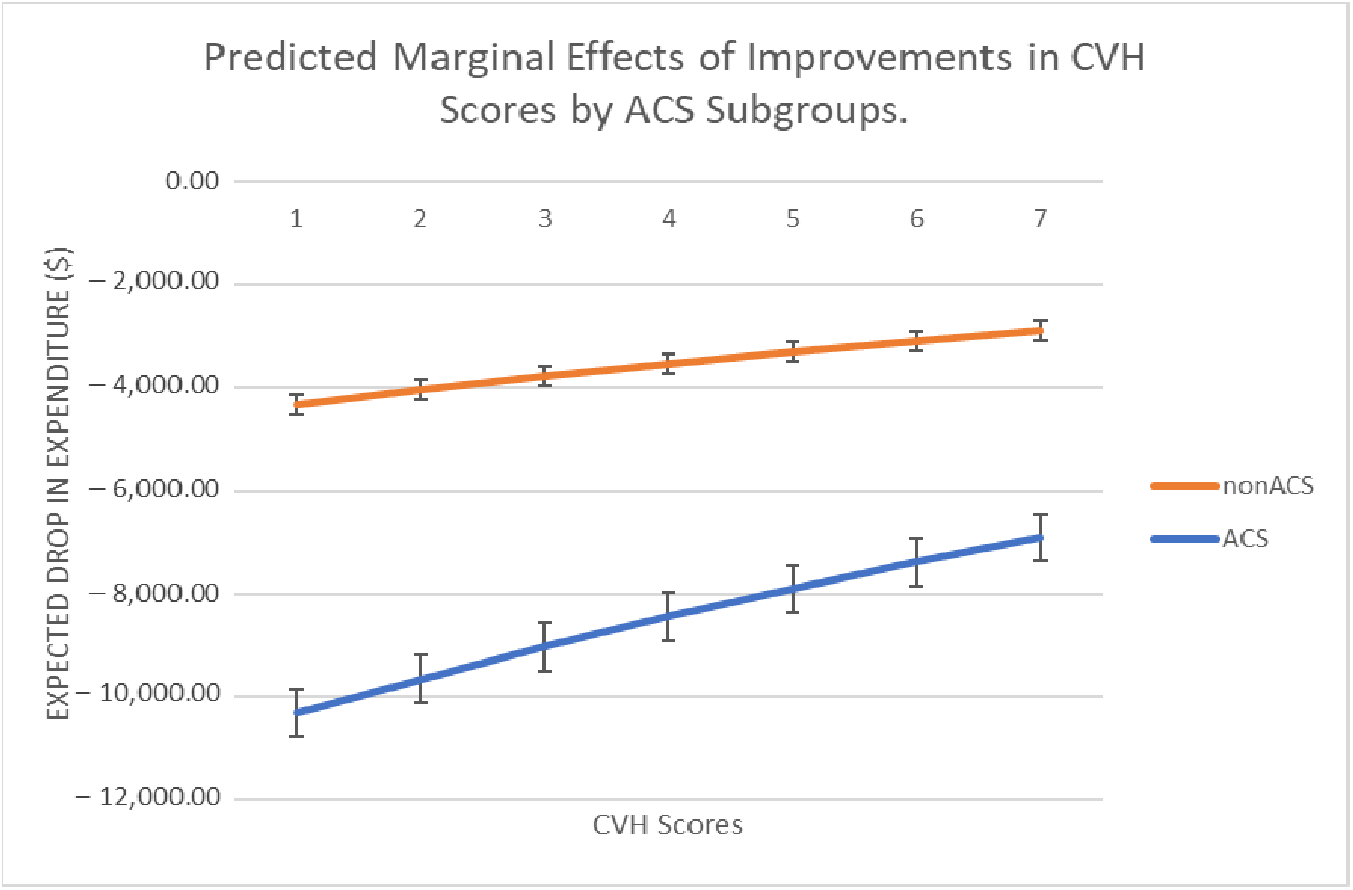
Marginal effects of improved CVH scores on savings.

### 3.4. Relative Variations in Expenditures of Life’s Simple 7 Components

Figure 3 below shows the variations in the impact on the level of expenditure for all CVH components. Meanwhile, achieving a normal BMI generates an annual variation of less than 6% and hypertension causes the highest variation in total healthcare expenditures (35.3%), closely followed by diabetes mellitus with 33% at p < 0.05. Inadequate physical activity and achieving normalized serum cholesterol levels had the least impact on expenditures, with −19% and 0.1%, respectively, at p < 0.05. These findings were consistent with the results of other researchers. For example, in Australia, scientists demonstrated that obesity, hypertension, and diabetes mellitus are predictors of higher pharmaceutical expenditure among persons with or at risk of CVD [18].

**Figure 3.**
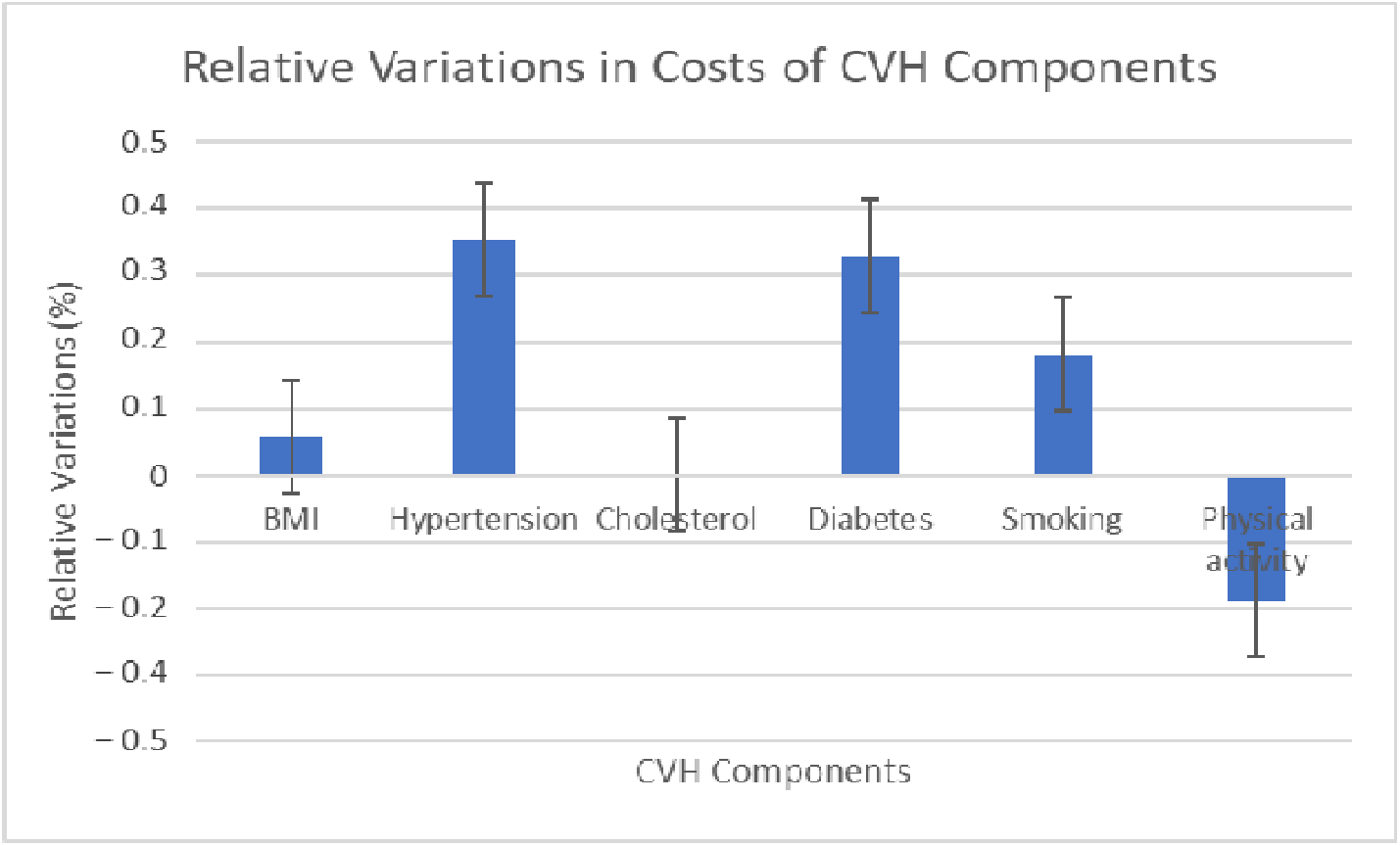
Bar graph representation of relative variations in cost of components.

## 4. Discussion

This study may be the first to evaluate the contrast in marginal effects of improvement in CVH on the percentage reduction in healthcare expenditure in ACS and non-ACS sub-groups in the United States. In a nationally representative population, our cross-sectional study demonstrated that between 2008 and 2018, adults with established ACS spent significantly more on healthcare compared with those without ACS. According to Table 2, if someone is aged 40–64 and has not had an ACS event to improve their CVH from the poor to the ideal range, it would cut their annual healthcare expenditure by more than half, saving them approximately $35,000 a year on average. This example is indicative of the great potential savings from improving the CVH scores of people who have low CVH Scores and who have not had an ACS event.

We also found significantly higher total healthcare expenditure (the sum of expenditures on inpatient, outpatient, emergency room, prescription drugs, home health, and office-based healthcare visits) to be associated with poor CVH profiles among persons with and without established ACS in a nationally representative cohort, after accounting for underlying comorbid conditions. This underscores the importance of addressing cardiovascular risk factor prevention in the general population, especially because the proper management of LS7 components reduces the risk of ACS. This approach can potentially lead to reduced healthcare spending or the redirection of existing healthcare spending to other healthcare needs. These findings are comparable to the findings of [23], in which they reported the impact of modifiable risk factors (which are essentially LS7 or CVH components) on average annual pharmaceutical expenditure among those with Atherosclerotic Cardiovascular Disease (ASCVD) in 2012–2013. They showed that overall costs increased with worsening CVH scores.

Some studies have demonstrated an association between comorbidity and healthcare costs [20]. In our study, increases in comorbidity levels based on Charlson’s comorbidity Index were positively associated with higher expenditures. The marginal increase in expenditure associated with worsening CVH scores was larger among those with ACS than those without ACS. The p-values of the results of the marginal effects of changes in expenditure levels as a function of increases in CVH management scores were highly significant at the 5% level. Many studies have found that current Cardiovascular risk factors are important drivers of future morbidity and mortality among individuals with established CVD in a dose-response fashion [7,25]. Therefore, effective secondary preventive measures for those who acquired the syndrome [17], through targeted educational endeavors, will reduce mortality and spending. Although studies within and outside the United States have attempted to estimate the incremental healthcare expenditures associated with individual cardiovascular risk factors among those without established CVD, no study has detailed the potential economic benefit of CVH management on total health expenses in both ACS subgroups. The consideration of people who have experienced ACS events matters since they currently carry the lion’s share of contributions to overall healthcare expenditures among people with chronic conditions [12].

After replacing the CVH metrics with individual CVH components in the regression, we captured the association between LS7 domains with total costs after controlling for all covariates. Diabetes and hypertension were found to impact costs the most. Meanwhile, achieving normalized blood cholesterol or trying to achieve moderate to vigorous physical activity was associated with the least total expenditure. These findings may find relevance in situations where understanding which of the LS7 components yields the most financial impact becomes critical for saving; for example, determining which two or three of the LS7 components would cost insurance or hospitals the most. Even though an association is not causation, where there is more smoke, there likely is more fire. These findings were consistent with the results of other researchers. For example, in Australia, Ademi et al. demonstrated that hypertension and diabetes mellitus are predictors of higher pharmaceutical expenditure among persons with or at risk of CVD [18].

Our study has important strengths and limitations. The large sample size increases the chance of finding statistically significant results and narrows down the confidence intervals of our findings. External validity was improved by how minority races or ethnicities were targeted and reweighted to make the results generalizable. Secondly, MEPS provides carefully executed interviews and study designs, which include multilevel verification of information collected from participants, for example double-checking costs and patient-reported information with primary-care physicians, hospitals, employers, and health insurance companies. The interpretation of our results should be performed with the following limitations in mind: The 3-digit nomenclature system (ICD-10-CM or ICD-9-CM codes) used to map medical conditions related to ACS could have underestimated the perceived prevalence of ACS. The reason for this is that not all health providers are able to accurately match symptom presentation with the diagnosis of ACS, again due to the less appropriate ICD coding of that specific diagnosis by coding personnel, causing a selection effect bias. Secondly, since cardiovascular risk factors (or CVH components) for determining CVH scores were self-reported, the actual national prevalence is likely underestimated. Given that MEPS does not consider costs associated with over-the-counter prescriptions, there might be an underestimation of the average expenditures on prescription medication, which constitute a subset of total healthcare expenditures. Finally, even though we chose covariates based on prior evidence that they are factors associated with higher or lower health expenditures, there may be unobserved confounders that could affect the expenditure habits of participants such as the type of health insurance purchased, the clinicians’ service rates, healthcare organizations policies, and individual patient behaviors that are major determinants of healthcare expenditures that are not assessed in this dissertation.

## 5. Conclusions

Overall, we found the annual impact of ACS on costs to be approximately $88,500. Modest changes in CVH metric scores generate savings in healthcare. Our study suggests that changes in behavioral measures and targeted modulation of health factors, especially normalizing blood pressure, blood glucose levels, body mass index, and smoking cessation may significantly reduce medical spending for ACS subgroups. While efforts to avoid ACS should be the first line of attack, when acquired, healthcare providers and insurance companies should adopt more aggressive secondary prevention strategies. CVH scores represent an important tool for secondary prevention that should be more widely used during cardiovascular rehabilitation (CR). CR is a comprehensive and multidisciplinary program that focuses on improving the health and well-being of individuals who have experienced a cardiovascular event. It addresses their physical, psychological, and social needs through exercise training, education on lifestyle behaviors (LS7), and counseling to manage psychological and emotional stress that come with ACS. Aligned with CR strategies, our study calls for the aggressive implementation of secondary CVD prevention among individuals affected by ACS.

## Data Availability

All data produced are available online at:
[https://meps.ahrq.gov/mepsweb/data_stats/download_data_files.jsp]

## Author Contributions

Conceptualization, A.E. and A.A.; methodology, A.E.; software, A.E.; validation, A.A., B.I., G.R., and N.C.B.; formal analysis, A.E.; investigation, A.E.; resources, A.E. and A.A.; data curation, A.E. and A.A.; writing—original draft preparation, A.E.; writing—review and editing, A.E., A.A., B.I., G.R., and N.C.B.; visualization, A.E. and N.C.B.; supervision, A.A.; project administration, G.R. All authors have read and agreed to the published version of the manuscript.

## Institutional Review Board Statement

The study was conducted in accordance with the Declaration of Helsinki, and approved by the Institutional Review Board (or Ethics Committee) of Florida International University by the Office of Research Integrity (Research Compliance, MARC 414) on April 26^th^, 2022. IRB-22-0170

## Informed Consent Statement

Informed consent was obtained from all subjects involved in the study

## Data Availability Statement

The data presented in this study are openly available in [https://meps.ahrq.gov/mepsweb/data_stats/download_data_files.jsp, accessed on 6 June 2020].

### Appendix A

**Table A1.**
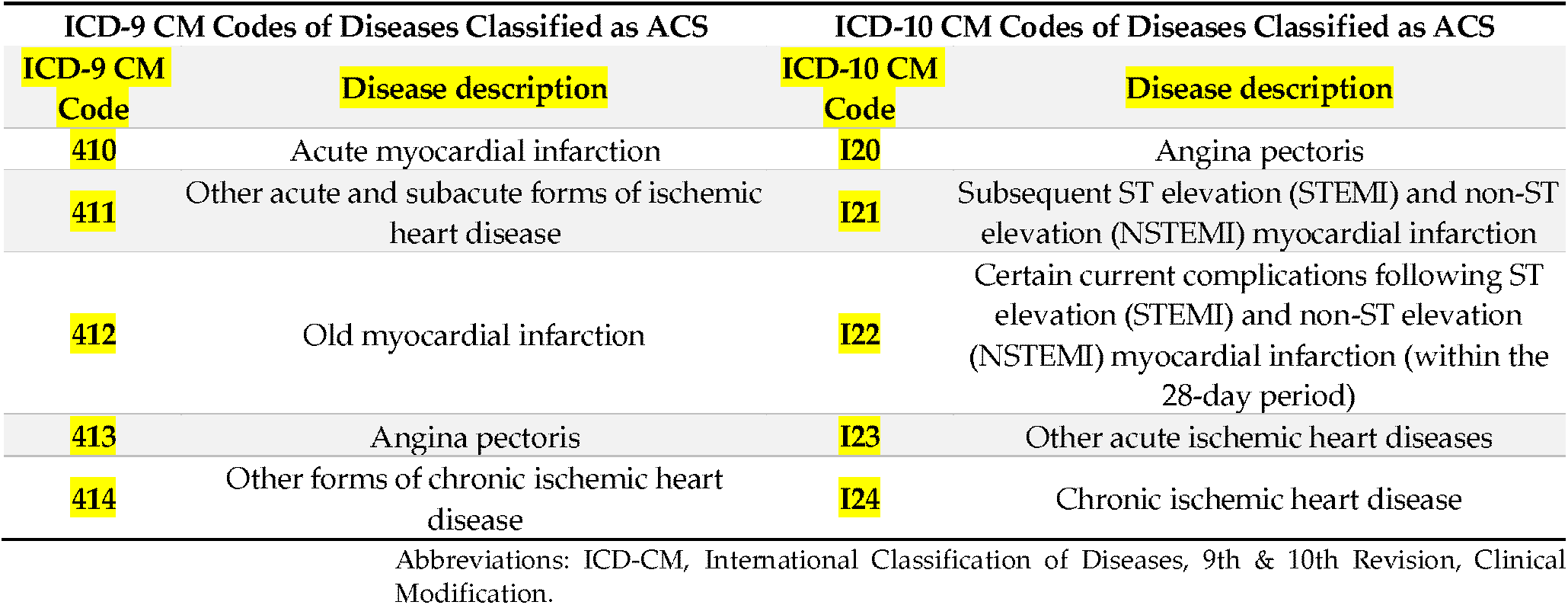
ICD-9 CM and ICD-10 CM codes of diseases classified as ACS.

**Table A2.**
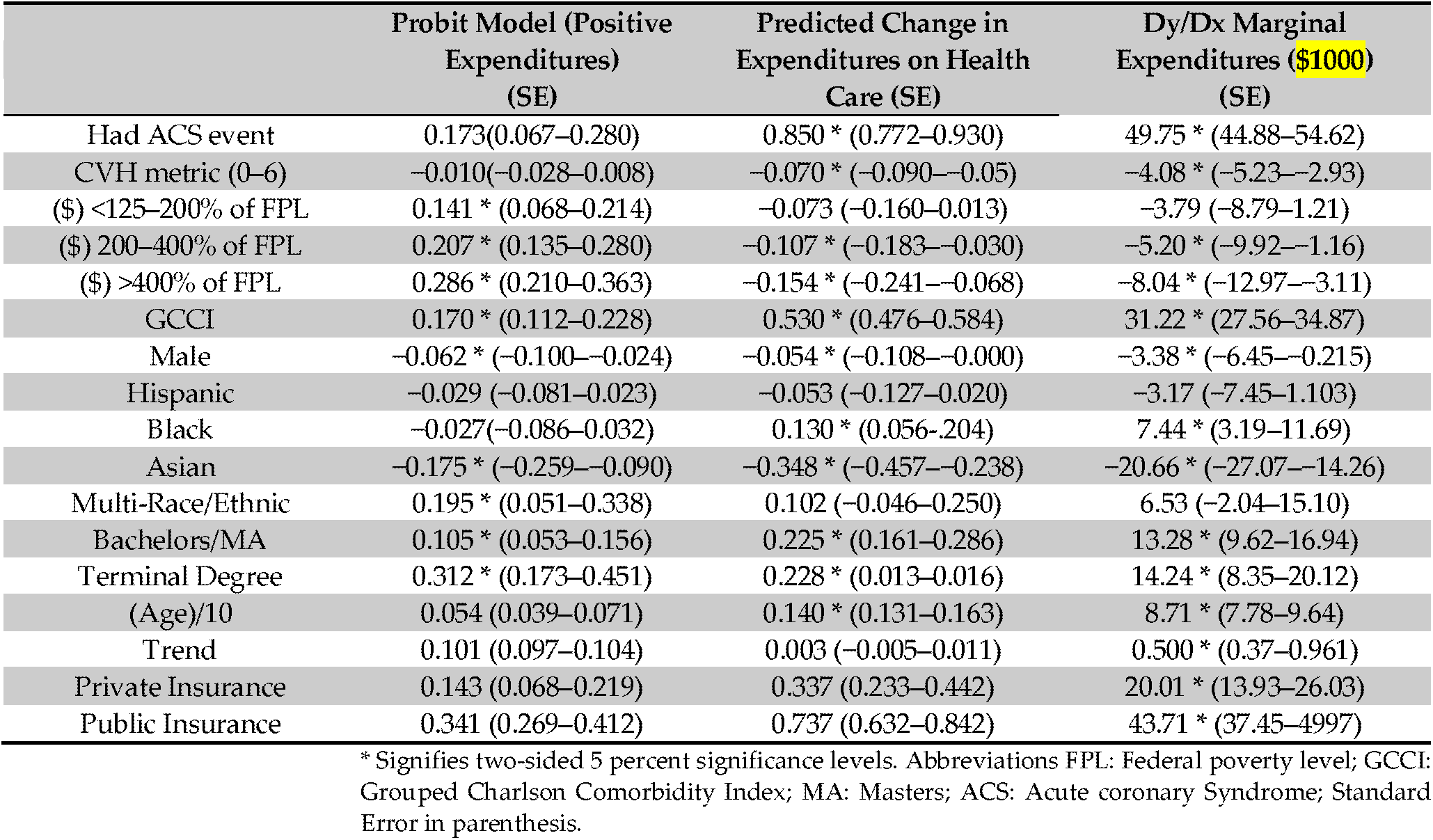
Two-stage model results for the prediction of expenditure based on CVH and ACS status for the years 2008–2018 using the MEPS dataset.

## Disclaimer/Publisher’s Note

The statements, opinions and data contained in all publications are solely those of the individual author(s) and contributor(s) and not of MDPI and/or the editor(s). MDPI and/or the editor(s) disclaim responsibility for any injury to people or property resulting from any ideas, methods, instructions or products referred to in the content.

